# Scaling COVID-19 rates with population size in the United States

**DOI:** 10.1101/2023.10.10.23296807

**Authors:** Austin R. Cruz, Brian J. Enquist, Joseph R. Burger

**Affiliations:** Department of Ecology and Evolutionary Biology, University of Arizona, Tucson, AZ, United States; Santa Fe Institute, Santa Fe, NM, United States; Department of Biology, University of Kentucky, Lexington, KY, United States

**Keywords:** Urban scaling, covid-19, epidemiology, scaling laws, allometry

## Abstract

We assessed Urban Scaling Theory using time-series data by quantifying allometric scaling relationships of coronavirus disease (COVID-19) cases, deaths, and demographic cohorts within and across three major variant waves of the pandemic (first, delta, omicron). Results indicate that with county-level population size in the United States, the burden of cases disproportionately impacted larger-sized counties. In contrast, the burden of deaths disproportionately impacted smaller counties, which may be partially due to a higher proportion of older adults who live in smaller counties. Future infectious disease burden across populations might be attenuated by applying Urban Scaling Theory to epidemiological efforts through identifying disease allometry and concomitant allocation of medical interventions.

## Introduction

The rapid global spread and persistence of the SARS-CoV-2 virus and disease (COVID-19) pandemic highlights the speed of interaction and magnitude of increasingly connected human populations in the twenty-first century. One of the key challenges to managing COVID-19 has been consistent and coordinated policy that controls the spatial and/or temporal dynamics of the virus in human populations (1, 2). Paramount to understanding how human infectious disease dynamics operate over space and time is an understanding of human population sizes and fluxes with respect to modern cities (3, 4).

Urban Scaling Theory (UST) is a general quantitative framework that provides predictions of how various physical, socio-economic, and biological attributes, including infectious disease, in modern cities change with population size as a power-law function (5, 6). Early analysis on the scaling of infectious disease comes from data on AIDS cases in the United States between 2002 and 2003, which showed a superlinear scaling exponent where larger cities demonstrated disproportionately more cases (6). Subsequent scaling analysis of AIDS cases in Brazil also showed superlinear scaling exponents in the years 2000 and 2010 (7). These results support the broader prediction that cities with larger population sizes are disproportionately more affected by disease than those with smaller population sizes likely due to a higher total number of contacts and speed of social interactions (8).

More recently, UST has been used to study the dynamics of COVID-19. Initial scaling research on COVID-19 in the United States demonstrated superlinear scaling of early pandemic case growth rates with city population size (9). Subsequent work in Brazil (10) and England and Wales (11) showed a temporal change in scaling exponents across the pandemic. At present, and given the United States initially represented a disproportionately large fraction of global COVID-19 cases and deaths (12, 13), we apply UST to COVID-19 cases, deaths, hospital beds (as a proxy for medical infrastructure), and younger and older demographic cohorts in the United States to characterize and predict how scaling parameters change with population size across the first two years (2020–2022) of the pandemic (see *Materials and Methods*).

## Results

Fig. 1 illustrates the time-dependent changes in scaling exponents (*β*, panel A) and normalization constants (*Y*_*0*_, panel B) for COVID-19 cases and deaths as a function of county population size within and across each variant. On the whole, *β* values for cases initially increase to a maximum for each variant (original: 1.3, 95% CI [1.18, 1.43], day 100; delta: 0.95, 95% CI [0.92, 0.98], day 567; omicron: 1.16, 95% CI [1.14, 1.18], day 720), and then slowly decrease thereafter. *β* values for deaths follow a similar pattern (original: 1.01, 95% CI [0.97, 1.06], day 200; delta: 0.65, 95% CI [0.58, 0.73], day 577; omicron: 0.86, 95% CI [0.82, 0.91], day 755). The analysis demonstrated a sublinear scaling between county population size and the county’s population aged 60 and older (*β* = 0.93 ± 0.0043 (SE); Fig. 2A), and linear scaling between county population size and the county’s population that is younger than 60 (*β* = 1.02 ± 0.0013 (SE); Fig. 2B). The scaling relationship between hospital intensive care unit (ICU) beds in a county and its population size was sublinear (*β* = 0.96 ± 0.014 (SE); Fig. 2C).

**Fig. 1.**
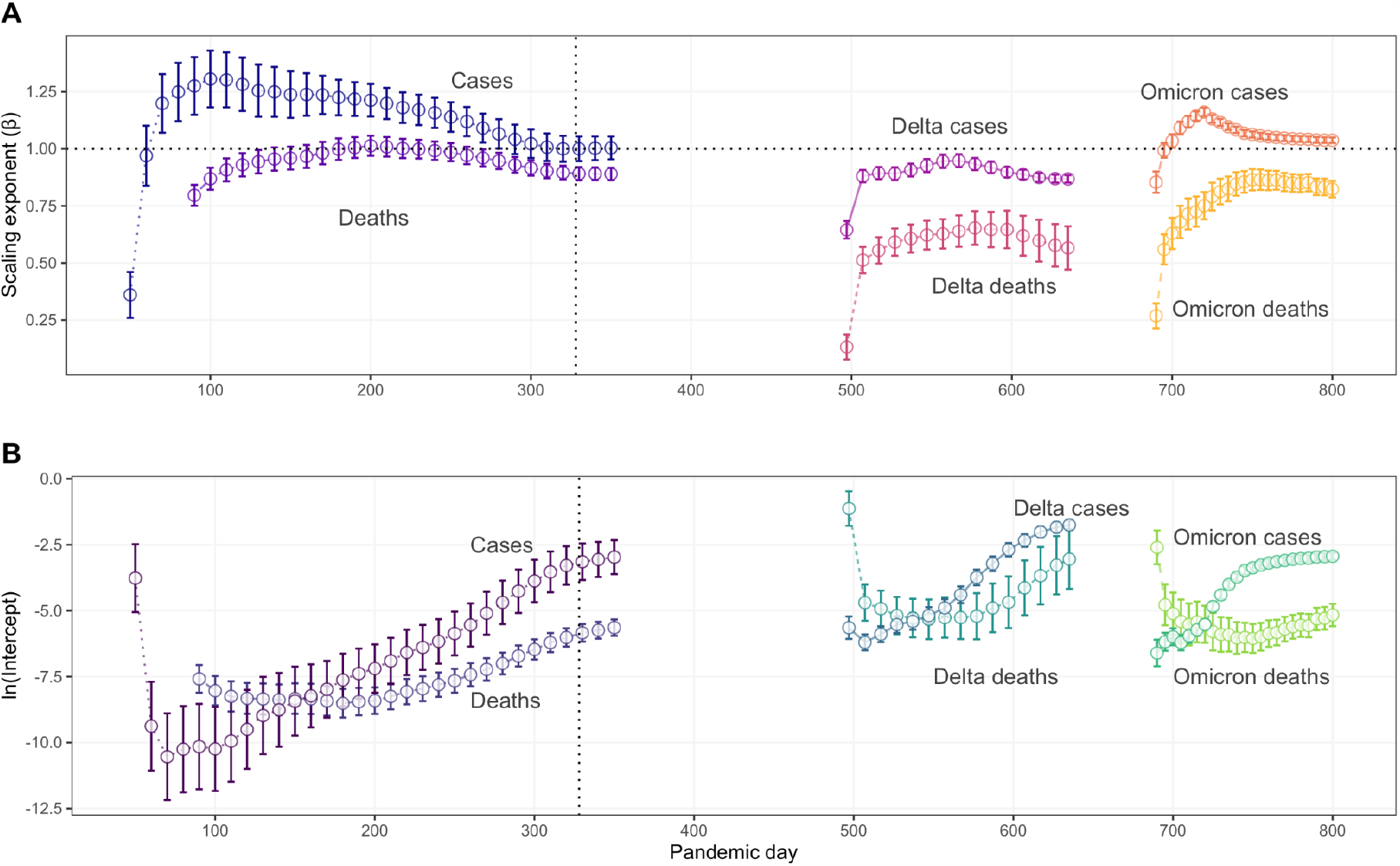
Allometric scaling behavior over time. A) Time-series change of scaling exponent (*β*) of COVID-19 cases and deaths. Dotted horizontal line indicates isometric scaling. B) Time-series change of normalization constant (*Y*_*0*_) of COVID-19 cases and deaths. Note: dashed vertical line in each panel indicates the start of vaccinations in the US (day 328).

**Fig. 2.**
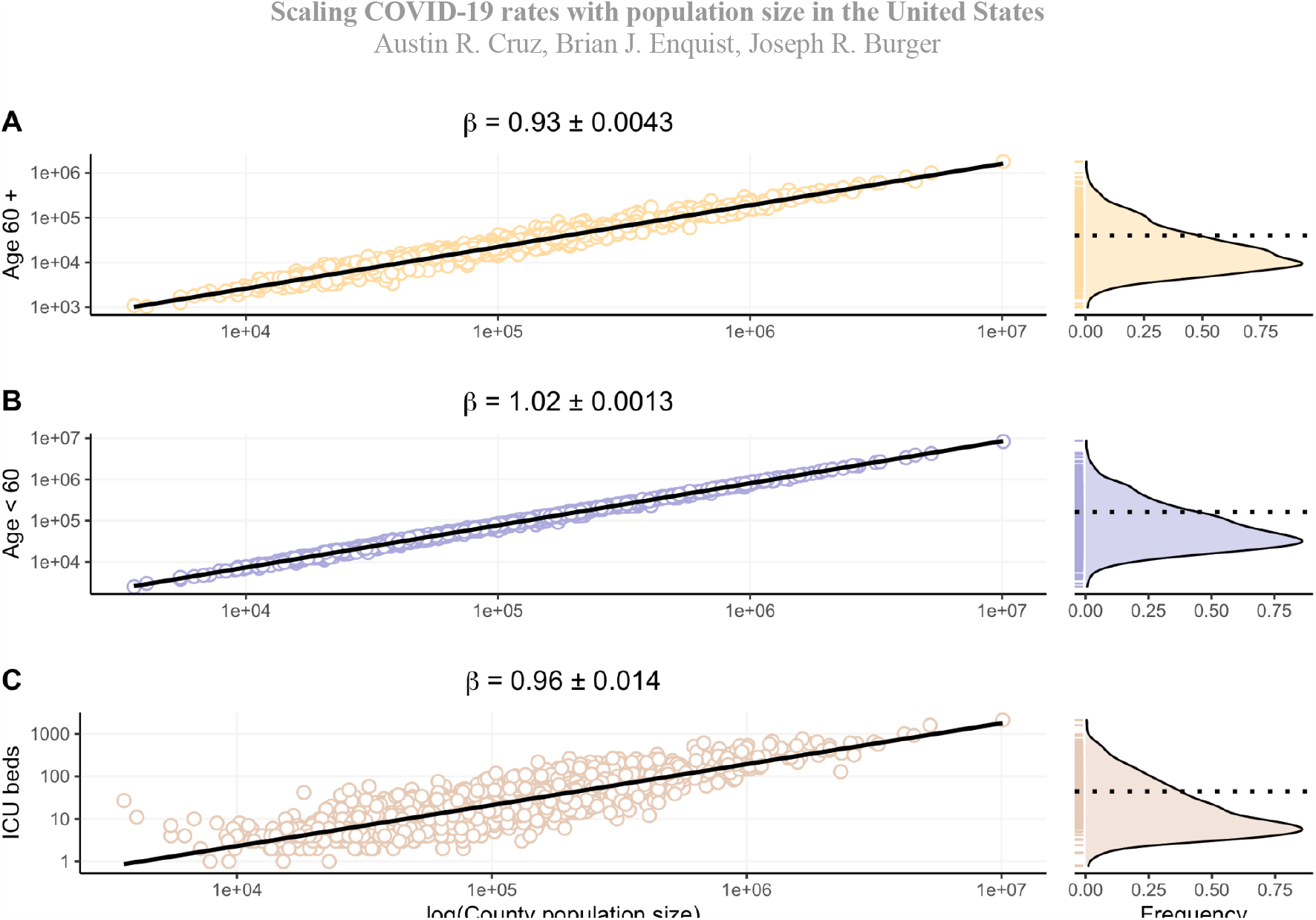
Allometric scaling of demographic cohorts and medical infrastructure. A) Smaller population counties have proportionally more older adults (β < 1) than larger counties. B) Younger populations are constant across counties (β ∼ 1). C) Smaller-county populations have disproportionately more IUC beds (β < 1). Frequency distributions and their mean values (dotted line) are shown to the right of each panel (A–C).

## Discussion

We show how allometric scaling relationships of COVID-19 cases, deaths, younger and older demographic cohorts, and medical infrastructure change with the United States’ county-level population size over the pandemic’s first, delta, and omicron waves. A key result is that the burden of cases disproportionately affects larger-sized counties, while the burden of deaths disproportionately impacts smaller-sized counties. This is demonstrated in the observation that the scaling exponent for cases is predominantly superlinear (> 1) for each variant, with a trend towards isometry (= 1). In contrast, the scaling exponent for deaths is consistently sublinear (< 1) for each variant. The exception to this observation is that delta cases remain sublinear over the analysis.

This larger pattern of superlinear cases and sublinear deaths may be due, in part, to a higher proportion of older adults who live in smaller-population counties. Policy and management strategies in the United States should consider how to scale medical care (e.g., hospital beds, medical professionals, antiviral medication, vaccinations) to address these differences. For example, differentially distributing medical and non-medical interventions to smaller counties may prevent the deaths of older individuals (14). Additional research may reveal differences in protective health measures in counties of different sizes, which may be related to, for example, past experience of disease burden (15). It is imperative that research in urban scaling delve deeper into the multiple mechanisms influencing epidemiological trajectories. Doing so can significantly amplify its relevance and effectiveness in ongoing and prospective epidemiological research and interventions.

## Materials and Methods

### Data

We used daily time-series data for COVID-19 cases and deaths, starting from January 22, 2020 (as day 1), from the Johns Hopkins University Center for Systems Science and Engineering COVID-19 Data Repository. These data included projected county-level population sizes in 2019 from 2010 US Census Bureau data for all counties (3142) in the United States. Data for county-level ICU hospital beds and the number of people aged 60 and older were obtained from Kaiser Health News (https://tinyurl.com/2s3pzfjp).

### Model

To assess the scaling relationship between COVID-19 cases and deaths, demographic cohorts, hospital ICU beds, and county-level population size in the United States, we used a power law function given by the model *Y*(*t*) = *Y*_*0*_ *N*(*t*)^*β*^, where *Y* is the given attribute for a county at time *t, N* is human population size as a measure of city size, *Y*_*0*_ is the normalization constant (intercept), and *β* is the exponent. Here, *β* reflects the rate at which the attribute changes with county population size, and is categorized among three classes of scaling behavior: sublinear (*β* < 1), linear or isometric (*β* = 1), and superlinear (*β* > 1). Parameters were estimated using the Ordinary Least Squares (OLS) regression method.

### Analysis

We grouped COVID-19 cases and deaths in the United States according to three major variant waves: first, delta, and omicron. The initial wave corresponds to days 1–350 in the analysis. Regression analyses for original variant cases and deaths start on day 90 and are reported every 10 days until day 350. The Centers for Disease Control and Prevention (CDC) reported that the COVID-19 delta variant became dominant in the United States on June 1, 2021 (day 497 in our analysis). From that day onwards, we assumed all new COVID-19 cases and deaths were attributed to “delta” in our analysis and calculated and reported new regression parameter estimates relative to the variant. The delta wave corresponds to days 497–635 in the analysis. Regression analyses for delta variant cases and deaths start on day 497 and are reported every 10 days until day 635. The CDC reported that during the week of Dec. 11–18, 2021 (days 690–697 in our analysis) the COVID-19 omicron variant became dominant in the US. From day 690 onwards, we assumed all new COVID-19 cases and deaths were attributed to “omicron” in our analysis and calculated and reported new regression parameter estimates relative to the variant. The omicron wave corresponds to days 690–800 in the analysis. Regression analyses for omicron variant cases and deaths start on day 690 and are reported every 5 days until day 800. Only counties that had one or more cases or deaths were included in the analysis for each variant.

## Data Availability

All data produced in the present work are contained in Zenodo repository (https://doi.org/10.5281/zenodo.8422680)

## Data availability

Code and associated data are available in Zenodo repository (https://doi.org/10.5281/zenodo.8422680).

## References

1. Kortessis, N., Simon, M. W., Barfield, M., Glass, G. E., Singer, B. H., & Holt, R. D. (2020). The interplay of movement and spatiotemporal variation in transmission degrades pandemic control. Proc. Natl. Acad. Sci. U.S.A., 117(48), 30104–30106.

2. Tuladhar, R., Grigolini, P., & Santamaria, F. (2022). The allometric propagation of COVID-19 is explained by human travel. Infectious Disease Modelling, 7(1), 122–133.

3. Burger, J. R., Weinberger, V. P., & Marquet, P. A. (2017). Extra-metabolic energy use and the rise in human hyper-density. Scientific Reports, 7(1), 1–5.

4. Burger, J. R., Okie, J. G., Hatton, I. A., Weinberger, V. P., Shrestha, M., Liedtke, K. J., Cruz, A. R., Feng, X., Hinojo-Hinojo, C., Kibria, A. S. M. G, Ernst, K. C., & Enquist, B. J. (2022). Global city densities: re-examining urban scaling theory. Frontiers in Conservation Science, 3, 879934.

5. Bettencourt, L. M. (2013). The origins of scaling in cities. Science, 340(6139), 1438–1441.

6. Bettencourt, L. M., Lobo, J., Helbing, D., Kühnert, C., & West, G. B. (2007). Growth, innovation, scaling, and the pace of life in cities. Proc. Natl. Acad. Sci. U.S.A., 104(17), 7301–7306.

7. Antonio, F. J., de Picoli Jr, S., Teixeira, J. J. V., & Mendes, R. D. S. (2014). Growth patterns and scaling laws governing AIDS epidemic in Brazilian cities. PLoS One, 9(10), e111015.

8. Schläpfer, M., Bettencourt, L. M., Grauwin, S., Raschke, M., Claxton, R., Smoreda, Z., … & Ratti, C. (2014). The scaling of human interactions with city size. Journal of the Royal Society Interface, 11(98), 20130789.

9. Stier, A. J., Berman, M. G., & Bettencourt, L. M. (2021). Early pandemic COVID-19 case growth rates increase with city size. npj Urban Sustain., 1(1), 1.

10. Ribeiro, H. V., Sunahara, A. S., Sutton, J., Perc, M., & Hanley, Q. S. (2020). City size and the spreading of COVID-19 in Brazil. PloS one, 15(9), e0239699.

11. Sutton, J., Shahtahmassebi, G., Ribeiro, H. V., & Hanley, Q. S. (2022). Population density and spreading of COVID-19 in England and Wales. PloS one, 17(3), e0261725.

12. Msemburi, W., Karlinsky, A., Knutson, V., Aleshin-Guendel, S., Chatterji, S., & Wakefield, J. (2023). The WHO estimates of excess mortality associated with the COVID-19 pandemic. Nature, 613(7942), 130–137.

13. Paglino, E., Lundberg, D.J., Zhou, Z., Wasserman, J.A., Raquib, R., Luck, A.N., Hempstead, K., Bor, J., Preston, S.H., Elo, I.T., & Stokes, A.C. (2023). Monthly excess mortality across counties in the United States during the COVID-19 pandemic, March 2020 to February 2022. Science Advances, 9(25), p.eadf9742.

14. Sasson, I. (2021). Age and COVID-19 mortality. Demographic Research, 44, 379–396.

15. Angelopoulos, K., Stewart, G., & Mancy, R. (2023). Local infectious disease experience influences vaccine refusal rates: a natural experiment. Proceedings of the Royal Society B, 290(1992), 20221986.

